# BURDEN OF COMMUNITY-ACQUIRED PNEUMONIA IN HUNGARY: A NATIONWIDE ANALYSIS OF INCIDENCE, HOSPITALIZATION RATES, AND MORTALITY BETWEEN 2016 AND 2020

**DOI:** 10.64898/2026.05.27.26354111

**Authors:** Zoltán Kiss, Zsófia Mészner, Andrea Kulcsár, Krisztina Bogos, Tamás Habon, Judit Moldvay, Zsolt Pápai-Székely, Lilla Tamási, Torzsa Péter, Zoltán Vokó, István Wittmann, Gergő Attila Molnár, György Rokszin, Valéria Kovács, Zsolt Abonyi-Tóth, Zsófia Barcza, Tamás G Szabó, Máté Várnai, Raymond Odhiambo, Andrea Berta, Miklós Darida, Ildikó Horváth, Krisztina Andrea Kovács, Natali Neuhauser, Botond Lakatos, Veronika Müller

## Abstract

**Background:** Community-acquired pneumonia (CAP) remains a major global health burden disproportionately affecting older adults and people with comorbidities, with Streptococcus pneumoniae as one of the leading bacterial causes in Europe. The Hungarian Occurrence and Burden of PnEumonia (Hungarian-HOPE) study examined the incidence, hospitalization rates, and mortality of CAP between 2016 and 2020 in Hungary.

**Methods:** The National Health Insurance Fund database was used to identify adult CAP patients (all-cause) based on ICD-10 codes J10–18. Outcomes included CAP incidence, 0–15-day hospitalization, and 0–30-day mortality after hospitalization, stratified by age, sex, and comorbidities (chronic obstructive pulmonary disease [COPD], asthma, cardiovascular disease [CVD], and type 1 and 2 diabetes [T1DM, T2DM]). Risk maps visualized relative risk gradients across population strata.

**Results:** During the pre-pandemic period (2016–2019), over 100,000 CAP cases and more than 50,000 hospitalizations were recorded annually. In 2020, recorded cases fell to approximately 98,000, while hospitalizations increased to 66,200. Hospitalization rates increased from 25.1% in 2016 to 29.1% in 2019, then increased to 43.1% in 2020. The 30-day mortality among hospitalized patients rose from 22.7% in 2016 to 23.6% in 2019. Incidence, hospitalization, and mortality all increased with age. Relative to healthy males aged 30–39 years, CAP risk escalated steeply in the ≥80 years cohort (incidence 5–15-fold; hospitalization >3-fold; mortality 11–24-fold) and was further amplified by COPD, CVD, or T2DM, with a lesser effect for T1DM.

**Conclusions:** The results highlight the substantial age- and comorbidity-driven CAP burden in Hungary and support prioritization of preventive strategies including pneumococcal vaccination for older adults and high-risk groups.

## INTRODUCTION

Community-acquired pneumonia (CAP) remains a leading cause of global morbidity and mortality, especially among older adults, immunocompromised patients, and those with chronic conditions. Beyond the acute phase, CAP is now recognized for its significant long-term consequences, including increased risks of cardiovascular (CV) events, respiratory impairment, and cognitive decline.^1,2^ *Streptococcus pneumoniae* is the primary bacterial cause of adult CAP in Europe. While meta-analyses historically attributed ∼25% of cases to this pathogen,^3^ recent data from the post-PCV10/13 era estimate that 18% of CAP cases are pneumococcal, with 49% caused by PCV13 serotypes.^4^ Pneumococcal disease (PD) spans a broad clinical spectrum from noninvasive forms, like pneumonia, to life-threatening invasive pneumococcal disease (IPD), including sepsis and meningitis.^5^

Globally, CAP incidence is estimated at 4,350 cases per 100,000 population.^6^ Regional burdens vary: the US reports ∼248 cases per 10,000 adults, while rates in Asia-Pacific settings can exceed 1,400 per 10,000 discharges.^6^ In Europe, incidence rises steeply with age, from 1–2 per 1,000 person-years in the general population to 14 per 1,000 in those aged ≥65 years.^7^ Men generally face higher rates and worse outcomes due to risk behaviors and comorbidities.^8,9^ Up to 50% of adult CAP episodes require hospitalization, especially among the elderly and those with comorbidities.^10,11^ Short-term mortality remains significant: 30-day mortality for hospitalized patients is typically 7–10%, but exceeds 20% in patients aged >80 years or those with multiple comorbid conditions.^12,13,14^ Risk factors for PD and CAP overlap significantly, with chronic conditions such as COPD, CVD, diabetes (T1DM/T2DM), asthma, and smoking substantially elevating risk.^15^ Quantitatively, the excess risk is clinically meaningful: for diabetes, the relative risk (RR) is ∼2.8 for pneumococcal pneumonia^16,17^ and 1.4–4.6 for IPD, with poor glycemic control further exacerbating CAP risk.^18^ Even higher risks are reported for chronic heart disease (RR 3.8) and COPD (RR 7.7) across multiple cohorts and meta-analyses.^19,20^

In Hungary, pneumococcal vaccination has been available since 2008, and the introduction of a mandatory infant pneumococcal national immunization program (PCV13) in 2014 substantially reduced the incidence of pneumococcal disease caused by vaccine-covered serotypes in children.^21^ Indirect protection through herd immunity has also been observed in adults, however, this effect appears limited in older age groups, particularly due to the persistence of certain serotypes, such as serotype 3, and ongoing serotype replacement (i.e. the emergence of non-vaccine serotypes).^22^ Despite these benefits, the burden of CAP in adults has not decreased proportionally in many settings, largely due to serotype replacement and the persistently high risk and low vaccination coverage in older adults with comorbidities.^23,24^ At the same time, pneumonia-related mortality has remained high or increased despite timely antimicrobial therapy, in the context of rising antimicrobial resistance driven by antibiotic overuse, which contributes to treatment failure and severe outcomes.^6^ Currently, pneumococcal vaccines for adults are recommended in Hungary for individuals aged ≥50 years and for those with underlying medical conditions, but vaccines are available only at out of pocket cost.^25^ Moreover, there is no adult National Immunization Program (NIP) or broad reimbursement scheme, which contributes to low vaccine uptake among elderly and high-risk populations.^26^

In addition, the lack of recent, comprehensive epidemiological data for CAP across the entire population has prevented supportive analyses for implementing an NIP targeting these vulnerable populations.

### Objective

To address this evidence gap, the Hungarian-HOPE study assessed the burden of all-cause community-acquired pneumonia (CAP) in Hungary by estimating incidence, hospitalization, and mortality rates by age, sex, and comorbidity status between 2016 and 2020 using the nationwide NHIF database. It also quantified excess risk versus reference groups through age-, sex-, and comorbidity-stratified risk maps to support identification of high-risk populations and inform vaccination and prevention strategies.

## METHODS

### Data Sources

This nationwide, retrospective, observational study was conducted using the databases of the Hungarian National Health Insurance Fund (NHIF) and the Central Statistical Office (CSO). The NHIF database is a comprehensive resource of ICD-10 codes related to reimbursed prescription claims, in- and outpatient visits and medical procedures, covering nearly 100% of the Hungarian population. The CSO database collects cause-specific mortality data for all deceased Hungarian citizens on an annual basis. Population denominators and cause-specific mortality counts by calendar year, age group and sex were obtained from the CSO and used as a reference for calculating annual rates and performing age standardization.

### Study Population and Case Definitions

Our analysis included patients with CAP (all-cause) identified from the NHIF database. Diagnostic information is recorded according to the International Statistical Classification of Diseases and Related Health Problems, 10th Revision (ICD-10). Any healthcare encounter in which the following ICD-10 codes were recorded as primary or secondary diagnoses was classified as a pneumonia case: viral pneumonia (J10–J12); bacterial pneumonia (J13–J15, including that caused by *Streptococcus pneumoniae*); and other/unspecified pneumonia (J16–J18). Cases were captured across inpatient and outpatient specialist care, prescription redemption data, and CT/MR examinations during the study period (January 1, 2016, to December 31, 2020). Patients were included if they had at least one pneumonia-related ICD-10 code (J10–J18) in a given year, supported by antibiotic treatment, chest imaging within ±30 days, or death within 30 days, or if pneumonia was recorded during inpatient care; recurrent episodes were defined by a ≥60-day gap between diagnoses within the same year. The index date was defined as the first appearance of a qualifying pneumonia ICD-10 code during the identification period. Each patient was followed for up to 60 days post-index to capture hospitalizations, rehospitalizations, and mortality outcomes. After 60 days post-index, another pneumonia related ICD code was taken as a new pneumonia case. Hospitalization of pneumonia was defined as any cause of hospitalization within 0–15 days after the first pneumonia ICD code. Hospitalized mortality was recorded if death occurred within 30 days after hospitalization with pneumonia related ICD code (0–30 days) (mortality was all-cause mortality).

### Comorbidity Assessment and Risk Stratification

Underlying medical conditions associated with an increased risk of severe PD were identified based on diagnosis and procedure codes recorded in in- or outpatient care between January 1, 2013, and the incident PD infection (index date). The following comorbidities were included as covariates in the analysis: type 2 diabetes mellitus (T2DM), type 1 diabetes mellitus (T1DM), chronic pulmonary diseases (various forms of acute and chronic bronchitis, emphysema, other forms of COPD and bronchiectasis), asthma bronchiale and cardiovascular diseases (CVD, including angina pectoris, acute myocardial infarction [MI], congestive heart failure [CHF] and stroke) (methods reference is detailed in Supplementary material).

Patients were assigned to mutually exclusive subgroups representing the presence of one or multiple chronic conditions, alongside a reference cohort of healthy individuals without recorded DM, CVD or pulmonary disease. All analyses were stratified by sex, age group, and risk category, with the 30–39-year-old healthy male population serving as the reference group for relative risk calculations.

### Statistical Analysis

All data were anonymized prior to analysis.

Statistical analyses were primarily descriptive and performed at the cohort and subgroup levels, with baseline characteristics (age group at index, sex, risk factors and region of residence) summarized as counts and percentages.

The main endpoints were the incidence rates of CAP (all-cause), hospitalization rates within 0–30 days from diagnosis, and all-cause mortality within 0–30 days from the diagnosis (only for hospitalized persons). Results were reported overall and by cohorts and subgroups as annual rates for 2016–2020. Annual trends were evaluated for each endpoint from 2016 to 2020, and outcomes were stratified by age, sex and comorbidity status.

Risk maps were generated to visualize relative risk gradients for CAP, hospitalization, and short-term mortality across population strata for the year of 2019. For risk comparison purposes across subgroups, rate ratios (RRs) were calculated for each age-sex-comorbidity combination relative to the reference population of healthy 30–39-year-old males (except mortality, where reference group was healthy 40–49-year-old male cohort - as there were no deaths in the 30–39 cohort during the analysis). Estimation precision was characterized using 95% confidence intervals (CIs), derived via the robust sandwich estimator to account for repeated annual observations.

All analyses were performed using R version 4.3.2 (R Foundation for Statistical Computing, Vienna, Austria). A p-value of <0.05 was considered statistically significant.

The study protocol was approved by the Hungarian National Ethics Committee (IV/3049-3/2021/EKU).

## RESULTS

### Study Population

We identified a total of 629,783 newly diagnosed cases of CAP in Hungary between 2016 and 2020 in the NHIF database. The annual number of newly diagnosed cases decreased steadily throughout the study period, with a cumulative reduction of 31.8% from baseline. Age distribution analysis for 2019 showed that most patients diagnosed with CAP were older than 50 years, with the highest incidence observed in the 60–69-year age group (22,931 cases) as seen in **Table 1**. Annual patient numbers with CAP and any cases are presented in **Supplementary Tables 1A and 1B**; age-specific CAP rates are detailed in **Supplementary table 2**.

**Table 1.**
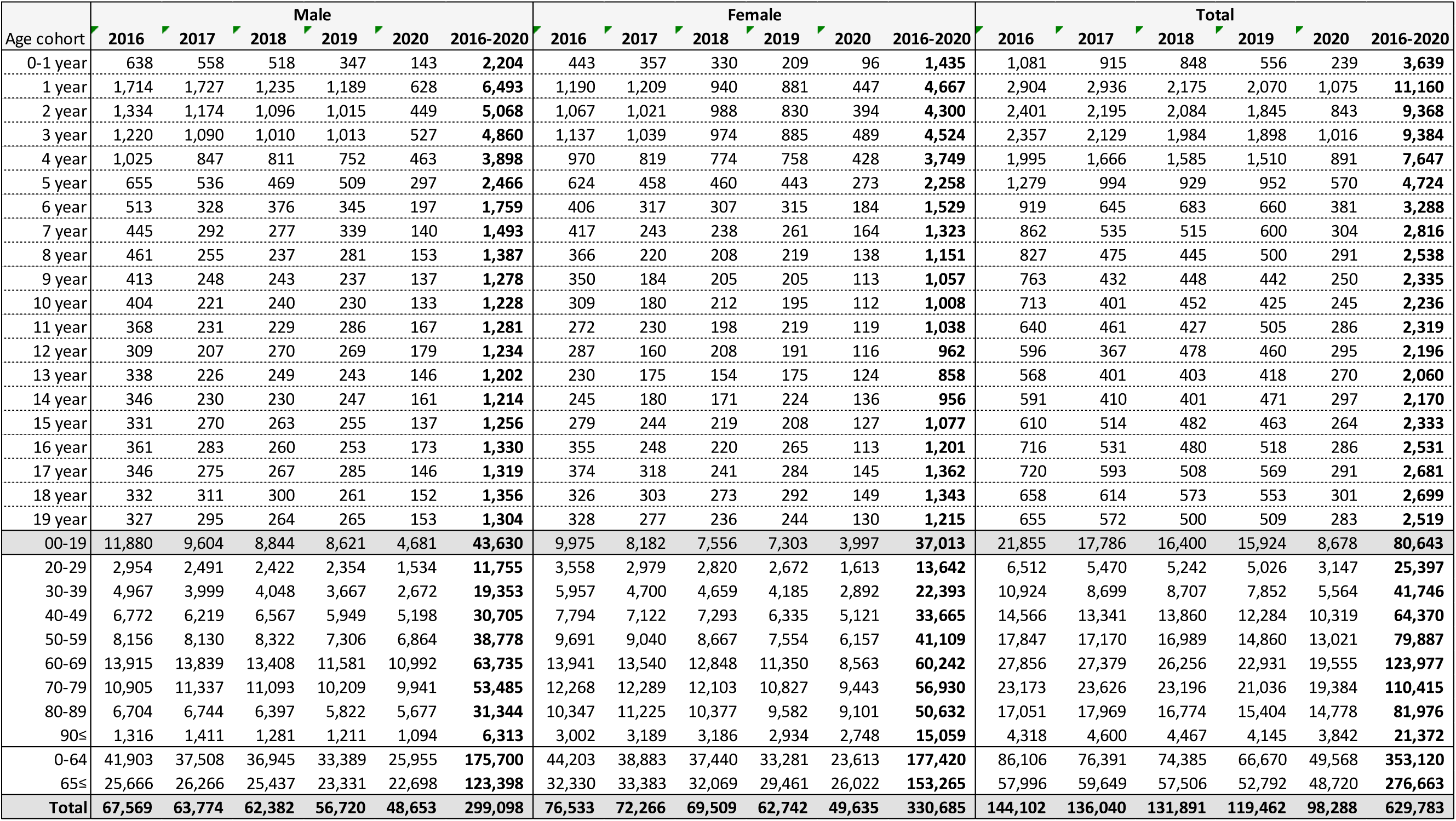
Annual number of diagnosed community-acquired pneumonia (CAP) cases by age and sex (2016–2020).

### Hospitalization Rates

The proportion of hospital admissions due to CAP increased steadily, rising from 25.1% in 2016 to 43.1% in 2020 (**Supplementary table 3A and 3B**). The male hospitalization rate was consistently higher than that of females and increased continuously until 2019 (**Figure 1**). In 2019, the hospitalization rate among pediatric and young adult patients varied between 51.5% and 11.5%, while lower rates were observed in the 20–49-year age groups (**Figure 2**). Among adults aged 50 and above, hospitalization rates increased progressively with age, reaching 19.1% (50–59 years), 27.9% (60–69 years), 37.8% (70–79 years), 51.2% (80–89 years) and 58.1% (≥90 years), respectively.

**Figure 1.**
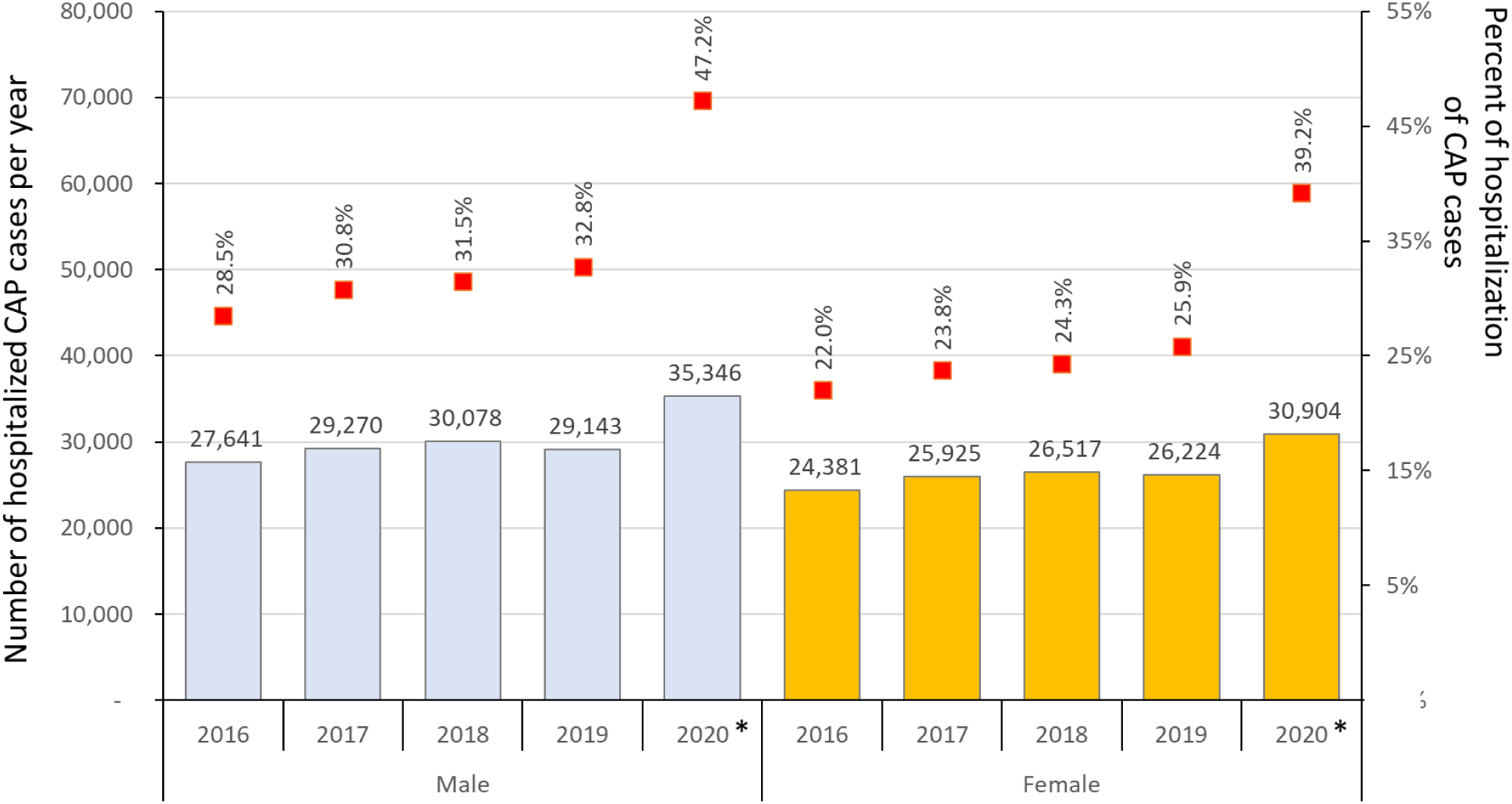
Annual number of hospitalized patients (colored bars) and hospitalization rates (red boxes) with CAP (0–15 days) by sex and study years between 2016–2020. *First year of the COVID-19 pandemic.

**Figure 2.**
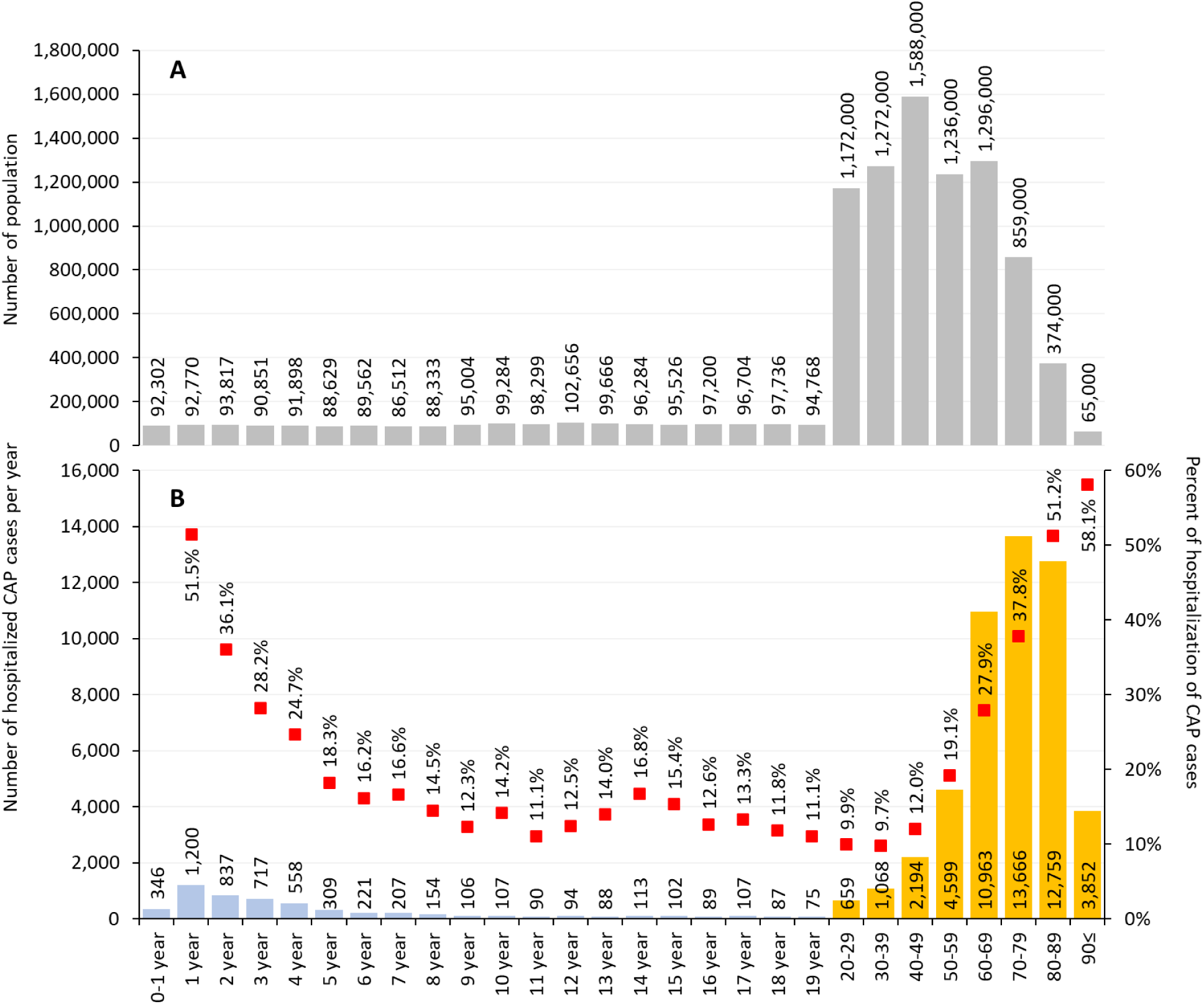
Annual number of population (A) and hospitalized patients (B) (colored bars – blue for 0–19 years by year and orange for >19-year-old cohorts) and hospitalization rates (red boxes) due to CAP by age group in 2019. Note that from the age of 20, ten-year age interval groups (orange bars) were formed.

### Thirty-Day Mortality Rates in Hospitalized Patients with CAP

CAP-related mortality within 30 days of hospitalization remained high throughout the study period of 2016–2020. The overall 30-day mortality rate among hospitalized patients increased significantly, rising from 22.7% in 2016 to 27.6% in 2020 from 11,788 yearly cases to 18,288 during this period (**Supplementary Table 4A and 4B)**. Males showed slightly higher overall deceased case counts and higher mortality rates, than females (**Figure 3**). Age-specific analysis demonstrated a clear age-gradient in 30-day mortality among hospitalized CAP patients, increasing steadily with advancing age and generally remaining higher in males. In patients younger than 50 years, mortality ranged from 0.8% to 13.3% in males and from 0.4% to 8.8% in females. Mortality then increased to 39.6% and 40.0% in females and males aged ≥90, respectively (**Figure 4**).

**Figure 3.**
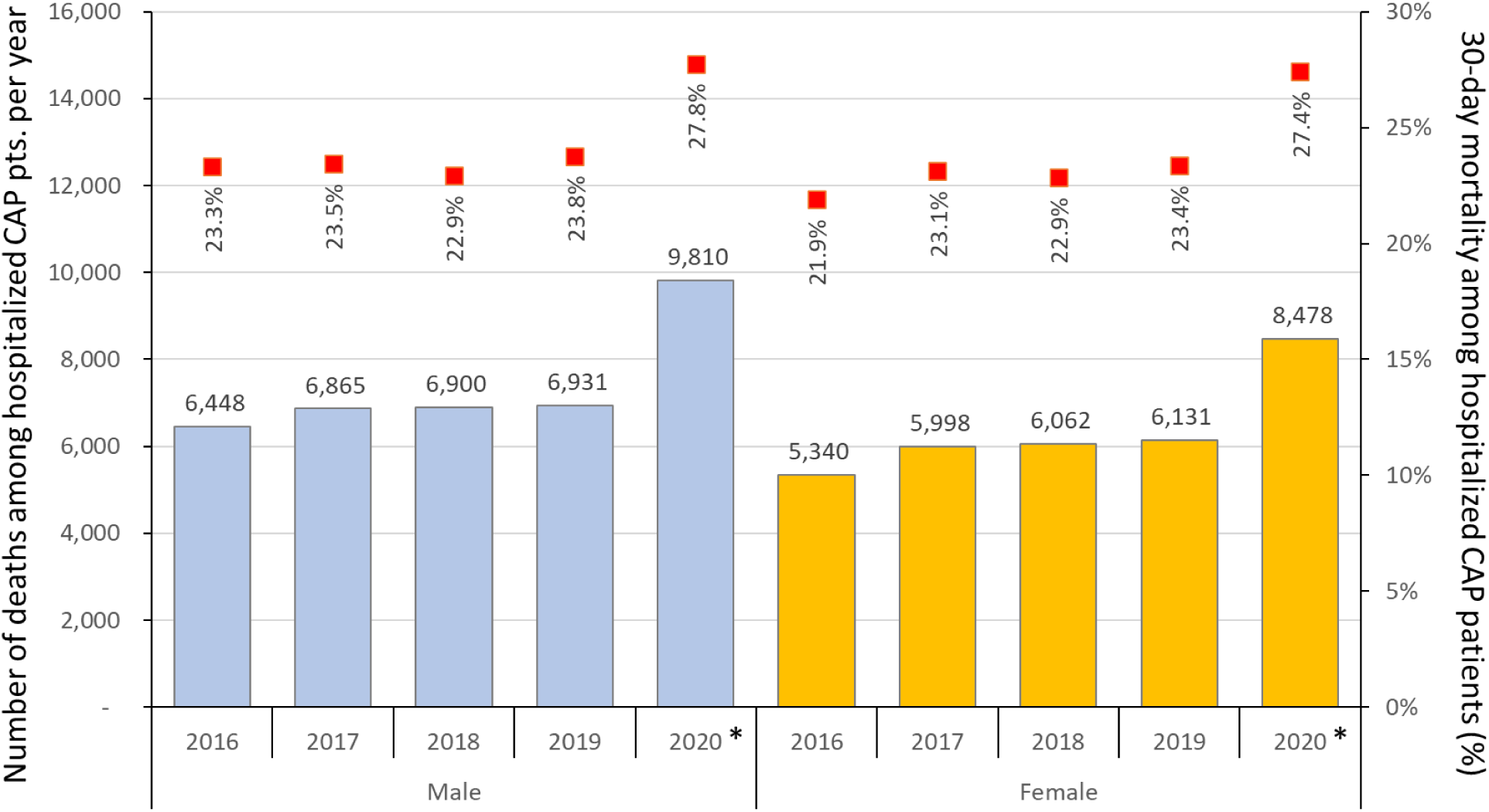
Annual number of deceased patients and mortality rates (0–30 day) among patients hospitalized due to CAP, 2016–2020. *First year of the COVID-19 pandemic.

**Figure 4.**
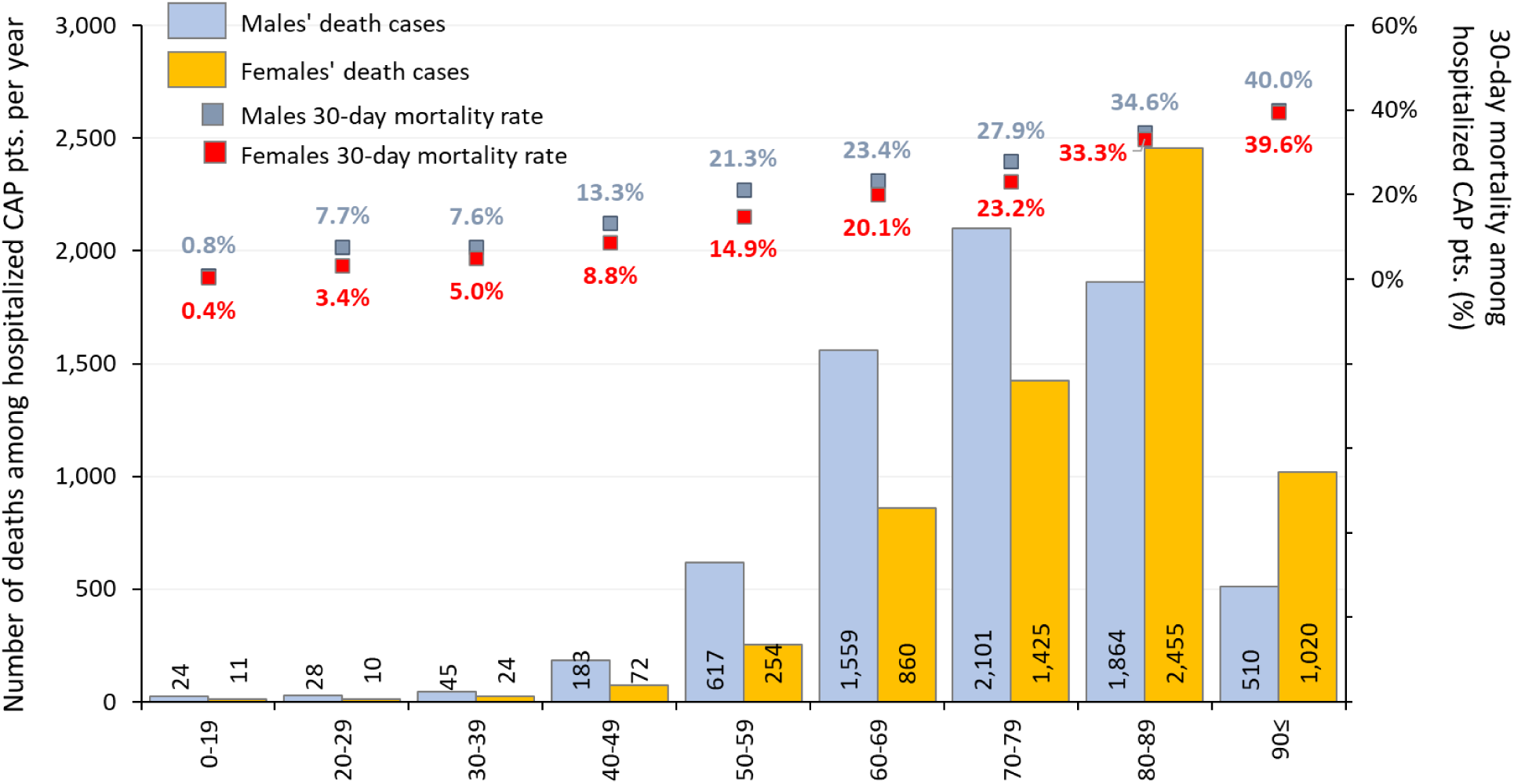
Annual number of deceased patients (colored bars) and mortality rates (30-day) (boxes) among patients hospitalized due to CAP by sex and age group in 2019, the last year before the COVID-19 pandemic.

### Risk Stratification by Age and Comorbidity

Based on risk estimates from 2019, the incidence of CAP increased steeply with advancing age, with male and female patients older than 80 years experiencing more than eightfold and fivefold risks, respectively, compared to the 30–39-year-old reference population. Chronic comorbidities were associated with a substantially elevated risk of CAP. The strongest independent risk factors were COPD and CVD: compared with the reference group without chronic disease, COPD was associated with a 9–10-fold increase in risk, while CVD was associated with a 7–9-fold increase in risk in the 40–49 age cohort (**Table 2**).

**Table 2.**
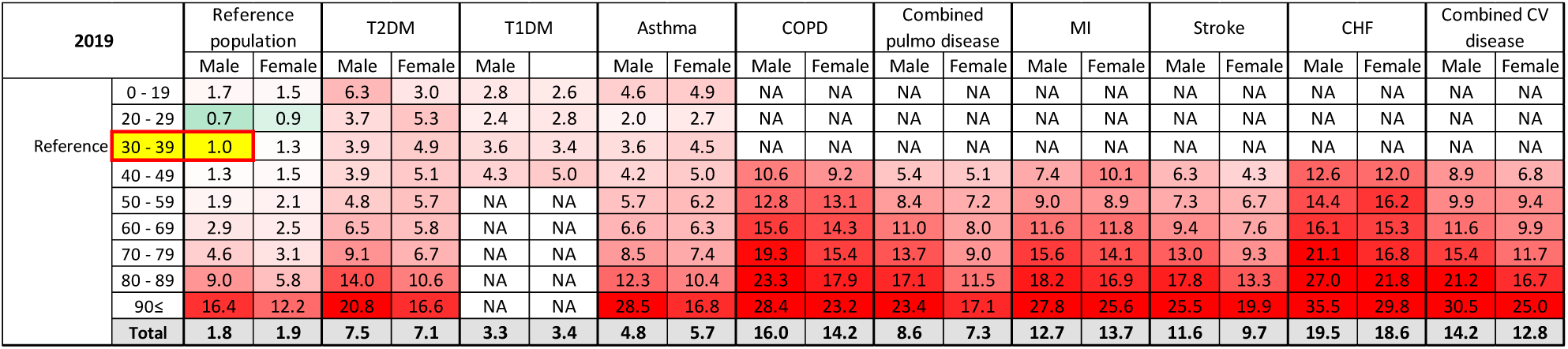
Heatmap showing relative *risk of community-acquired pneumonia (CAP)* in 2019 by comorbidity status, sex, and age group, compared with the reference population (healthy males aged *30–39 years*).

CAP-related hospitalization rates increased substantially with advancing age and the presence of comorbidities. Compared with healthy adults aged 30–39 years, the risk of CAP-related hospitalization was approximately twofold higher in patients aged 60–69 years, and more than threefold higher in those aged over 80. The presence of COPD, stroke or CHF increased the likelihood of hospitalization notably, already in the 40–59-year-old population (**Table 3**).

**Table 3.**
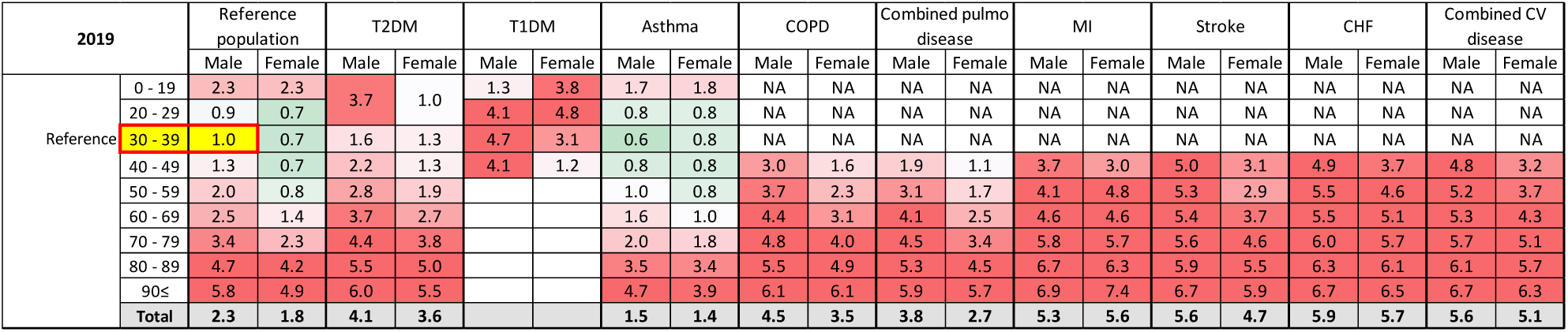
Heatmap showing relative *risk of hospitalization* within 0–15 days after community-acquired pneumonia (CAP) diagnosis in 2019, stratified by comorbidity status, sex, and age group, compared with the reference population (healthy males *aged 30–39 years*).

Mortality risk associated with CAP increased even more sharply with age: patients aged 70–79 years had a 4–8-fold higher risk of mortality compared with the reference group, and those aged 80 years or older exhibited more than an 11-fold increase. The presence of COPD, CHF or DM further elevated CAP-related mortality risk, especially for middle aged patients, with the highest relative risk observed among elderly male patients with CV comorbidities (**Table 4**).

**Table 4.**
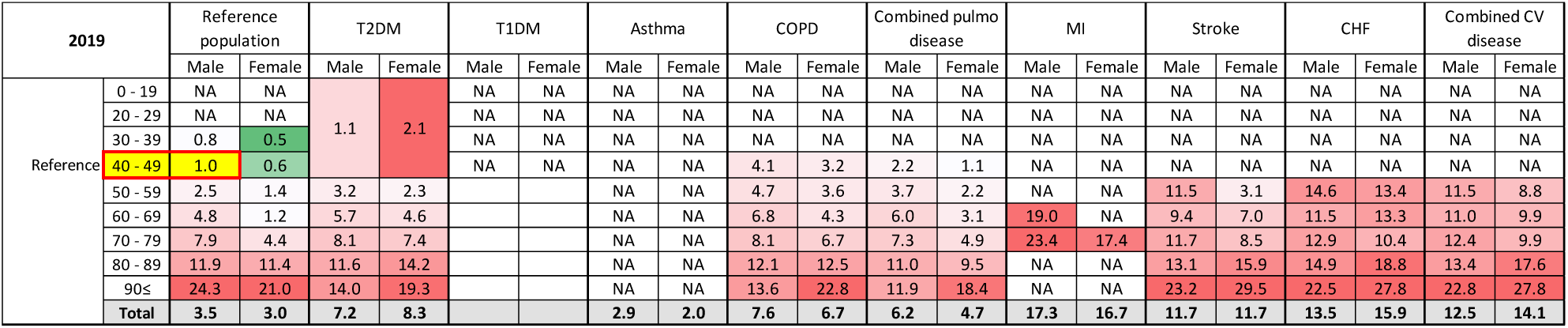
Relative *risk of 30-day all-cause mortality* after community-acquired pneumonia (CAP) diagnosis in 2019, stratified by comorbidity status and age group, compared with the reference population (healthy males *aged 40–49 years*).

Details regarding the chronic conditions considered for access risk calculations are shown in Supplementary Tables 5).

## DISCUSSION

### Incidence

The nationwide, retrospective Hungarian-HOPE study is the first comprehensive analysis of the epidemiology of CAP in Hungary between 2016 and 2020. The results show that CAP constitutes a substantial burden in Hungary, with over 100,000 cases/year and more than 50,000 annual hospitalizations following diagnosis during the pre-pandemic years (2016–2019 period and 0–30-day hospitalization rate). Although we observed a decreasing tendency in annual case numbers, our study period was not long enough to determine definite trends. The drop in the number of newly registered cases in the first year of the COVID-19 pandemic (from 119,462 in 2019 to 98,288 in 2020) compared to the preceding years is unlikely to reflect a true epidemiological improvement; it most likely resulted from social distancing, mask use, reduced mobility, and lower willingness to seek medical advice.^27^ Similar drops in non-COVID pneumonia presentations in 2020 have been reported by other European studies,^28,29^ which supports the assumption that apparent decline in annual case counts reflects behavioral and healthcare service changes during the pandemic.

The incidence of CAP showed strong associations with age, with steep increases after middle age, which is in line with European population-based data. The reasons behind this include the aging of the immune system, the higher prevalence of chronic comorbid diseases, and reduced ability to defend against respiratory infections. In Germany, the incidence of adult CAP was approximately fourfold higher in older, than in younger adults, and was strongly amplified by at-risk and high-risk conditions.^11^ A recent systematic literature review highlighted wide variations in CAP incidence across Europe, with the highest burden observed in advanced age groups and those with comorbidities, as evidenced by elevated rates in inpatient/outpatient settings primarily among the elderly. The review noted exponentially increasing hospitalization and mortality rates with age, aligning with prior European data (e.g., 2–10-fold higher in ≥85 vs. 50–64 years in Central Eastern European countries). *Streptococcus pneumoniae* predominated etiology in older adults, driving economic costs mainly from prolonged inpatient stays.^30^

### Hospitalization

We identified more than 50,000 cases of CAP-related hospitalizations each study year between 2016 and 2019 (within 0–15 days of an index diagnosis). The proportion of cases requiring hospitalization increased from 25.1% in 2016 to 29.1% in 2019. In 2020, the higher rate of hospitalization (43.1% may have been a result of the second wave of the COVID-19 pandemic leading to more severe cases of viral pneumonia and lower admission threshold. The incidence of hospitalization for adult CAP has repeatedly been shown to increase over time and to be concentrated in older age groups.^31^ For example, in England, age-standardized CAP admissions increased by 34% between 1997 and 2005 and continued to show upward pressure into the 2010s.^32^ A retrospective study from France found age-dependent increases in the incidence of adult CAP hospitalizations between 2013 and 2019, with patients aged ≥65 years accounting for 75% of hospitalization episodes.^33^ The increase in CAP hospitalization rates may be attributed to the ageing of the population and the resulting increase in the prevalence of chronic conditions; a more cautious, risk-averse approach of hospital staff; and the slight increase in the severity of CAP.^31^

Hospitalization rates in our study significantly varied with age. Patients younger than 20 years were mainly admitted for preventive purposes (∼22.5%), nevertheless, in pediatric populations, data indicate a particularly high burden of severe disease in the youngest age groups; for example, Jain et al. found that children aged 0–4 years exhibited markedly higher hospitalization rates and severe outcomes due to community-acquired pneumonia compared with older children. ^34^ There was a steep age-dependent increase in the adult population: only 9.9% of patients aged 21–30 years required hospital admission (2019), whereas hospitalization rates of CAP exceeded 50% among patients aged ≥80 years. This age-dependence is consistent with German data showing that the incidence of CAP hospitalization is nearly 1.5-fold higher among patients aged ≥60 years compared to younger cohort^11^ and other European datasets indicating that older patients account for the majority of inpatient CAP burden.^33^ The 2020 increase in CAP hospitalization rates, despite fewer recorded cases, parallels European reports from the pandemic period showing that non-COVID pneumonia admissions declined in absolute number, but were disproportionately concentrated among more severe cases.^33^ Among children and adolescents, higher hospitalization rates are common in routine practice and may in part reflect precautionary admission thresholds; this interpretation is supported by pediatric respiratory literature describing case-mix shifts during periods of non-pharmaceutical interventions.^28^

### Mortality

Thirty-day mortality rates of patients hospitalized with CAP increased from 22.7% in 2016 to 23.6% in 2019 and exceeded 27% in 2020. This phenomenon may be explained by the increasing mean age and comorbidity burden among admitted patients, the increasing proportion of severe cases requiring hospitalization, potential changes in pathogen virulence/profile, and a higher risk of hospital-acquired infections. Mortality also exhibited a clear age gradient, with rates of 0.6%, ∼7%, ∼23%, and >40% in patients aged <20, 20–40, 60–69, and >80 years, respectively. An age-limit of 65 years is also used for the estimation of 30-day mortality of CAP, and included into the CURB-65 score aiming to help deciding the need of hospitalization of cases with CAP. In the present study, mortality rates were also approx. twice as high for patients aged 65+ as that of their younger counterparts (Suppl. Table 4). European claims-based analyses corroborate substantial short-term mortality after CAP hospitalization, with a pronounced age gradient. In Germany, reported mortality was 18.5% in-hospital, 22.9% at 30 days, and 44.5% at 1 year, with mortality risks more than twofold higher in older versus younger adults.^11^ Based on European time-trend analyses and pandemic-era evaluations, increases in short-term mortality over time may reflect an older and more comorbid admitted population, delayed presentation, shifts in the microbial landscape, and periods of health-system strain, which are factors discussed in European time-trend analyses and pandemic-era evaluations.^31,29^ The comparatively high mortality observed in the 20-40-year group in our dataset warrants further consideration and may indicate selective hospitalization of more severe phenotypes (e.g., immunocompromised) and/or delayed care seeking, rather than a uniform increase in risk among all younger adults. Similar signals have been described in administrative datasets, where hospitalized ‘younger’ CAP patients can still experience 30-day mortality rates of ∼10%.^11^

The marked increase in 30-day mortality observed in 2020 likely also reflects the impact of the COVID-19 pandemic. During the pandemic, hospitalized pneumonia cohorts increasingly included patients with SARS-CoV-2 infection, either as primary viral pneumonia or as bacterial superinfection, both associated with substantially higher short-term mortality. In addition, healthcare system strain, delayed presentation, and reduced access to timely care during pandemic peaks have been shown to adversely affect outcomes in acute respiratory infections across Europe. Pandemic-era analyses consistently report increased in-hospital and 30-day mortality among pneumonia patients in 2020 compared with pre-pandemic years.^35,36^

### Impact of Age and Comorbidities

To examine and quantify the impact of age and comorbidities on the risk of CAP and its associated mortality compared to healthy reference groups, risk maps stratified by age, sex, and comorbidity were created for the Hungarian adult population. Older patients and those with COPD, CVD, or T2DM (and T1DM too, nevertheless, it was lower than T2DM) exhibited marked excess risks of CAP incidence and mortality compared to the healthy 30–39-year-old population. The impact of older age on relative risks was particularly pronounced in patients with COPD or CVD: compared with healthy males of the same age, the risk of CAP was 6–9-fold higher in those aged 30–39, while it was 15–20-fold higher in those aged >70 years. This is consistent with European evidence showing that chronic cardiometabolic and pulmonary conditions increase the risk of CAP and are associated with poorer outcomes.^15,37^ Population-based data from Germany indicate that CAP rates increase substantially in people with at-risk/high-risk comorbidities, beyond the effect of age alone.^11^ When set against EU literature, the relative-risk patterns in our maps appear directionally concordant with findings from other European settings. Across European cohorts, COPD/chronic respiratory disease typically confers an approximately 2–3-fold increase in adult CAP risk. For example, a recent Catalonian cohort study reported an adjusted hazard ratio of ∼2.66 for pneumococcal pneumonia (PP) among adults aged ≥50 years,^38^ in line with earlier European reports that identify chronic lung disease among the strongest determinants of CAP.^11,15^ For T2DM, European estimates generally suggest a more modest excess (roughly 1.2-1.8-fold for CAP/PP), and the increases seen on the Hungarian-HOPE risk maps fall within this range.^15,38,39^ Similarly, chronic heart disease is commonly associated with a ∼1.3–2.0-fold increase in CAP/PP risk in European studies, again mirroring the magnitude observed in Hungary.^11,15,38^ Finally, the graded increase with multimorbidity in our maps (combined pulmonary conditions), suggesting additive and potentially synergistic effects, is strongly supported by Catalonian data, where the risk of PP increased from HR 1.76 in those with one condition to HR 8.70 in those with ≥5 conditions.^38^ Overall, age-, disease-, and comorbidity-stratified excess risks in our study are both directionally and quantitatively consistent with evidence from the European Union, supporting the external validity of our risk maps and strengthening their implications for targeted prevention policy.

### Prevention strategies

As pneumococcus remains as one of the leading bacterial cause of adult CAP in Europe,^10^ and the risk is concentrated in older adults and people with comorbidities, pneumococcal vaccination should be a cornerstone of prevention policy.^37^ The CAPiTA randomized trial in adults aged ≥65 years showed that 13-valent pneumococcal conjugate vaccine (PCV13) reduced vaccine-type CAP by approximately 46% and vaccine-type invasive pneumococcal disease by about 75%, providing clear proof that conjugate vaccination can prevent serious pneumococcal outcomes, even if effects on all-cause CAP are limited.^40^ Recent European policy reviews continue to document low adult pneumococcal vaccine uptake and substantial heterogeneity in national recommendations; reported coverage in adults aged ≥65 years varies widely across countries (roughly 1–70%), with average uptake remaining below 20% in many settings.^41,42^ Experience from countries that have implemented comprehensive National Immunization Programs (NIPs) and structured delivery approaches (e.g., standing orders, reimbursement, pharmacy-based vaccination, and reminder/recall systems) suggests that improving access and system-level prompts can raise coverage and is associated with reductions in PD-related hospitalizations and deaths, supporting the rationale for strengthening risk-stratified adult immunization strategies.^22^ Across Europe, an increasing number of countries have introduced publicly funded pneumococcal vaccination programs for adults, employing age-based and/or risk-based strategies to improve vaccine uptake and reduce the burden and mortality associated with pneumococcal disease.

### Strengths and limitations

The main strengths of our study lie in the nationwide nature of the NHIF database, the uniform ICD-10 definitions, and the integrated hospitalization/mortality follow-up. Determining the etiology of CAP remains inherently challenging due to potential diagnostic misclassification and limitations in routine microbiological testing. Of note, the NHIF database did not allow for the reliable differentiation of PD etiology from all-cause CAP in the Hungarian-HOPE study; therefore, conclusions regarding the burden of PD and the rationale for preventive immunization were based on available reports on the proportion of PD among CAP cases and should be interpreted with caution. Additionally, the NHIF database does not capture individual-level information on pneumococcal vaccination status or key behavioral risk factors such as smoking, which may have resulted in residual confounding and limited the assessment of their contribution to CAP risk and outcomes.

Finally, the lack of age- and sex-standardized rates for all Hungarian-HOPE outcomes warrant further investigations in extended analyses of our data to allow for comparisons with reports from the European Union.

## CONCLUSIONS

The HOPE study is the first comprehensive analysis of the epidemiology of CAP in Hungary. The results show that the incidence, hospitalization, and mortality rates of CAP increase with age and the number of chronic conditions, which helps identify populations in which targeted prevention, especially vaccination including against *S. pneumoniae*, should be high public health priority.

## Data Availability

All data produced in the present work are contained in the manuscript as tables and supplementary material.

