## Supplemental Tables 1-4 for "BURDEN OF COMMUNITY-ACQUIRED PNEUMONIA IN HUNGARY: A NATIONWIDE ANALYSIS OF INCIDENCE, HOSPITALIZATION RATES, AND MORTALITY BETWEEN 2016 AND 2020"

**Supplementary Table 1A** Total annual number of patients with a recorded community-acquired pneumonia diagnosis, by study year, age group, and sex (2016-2020)

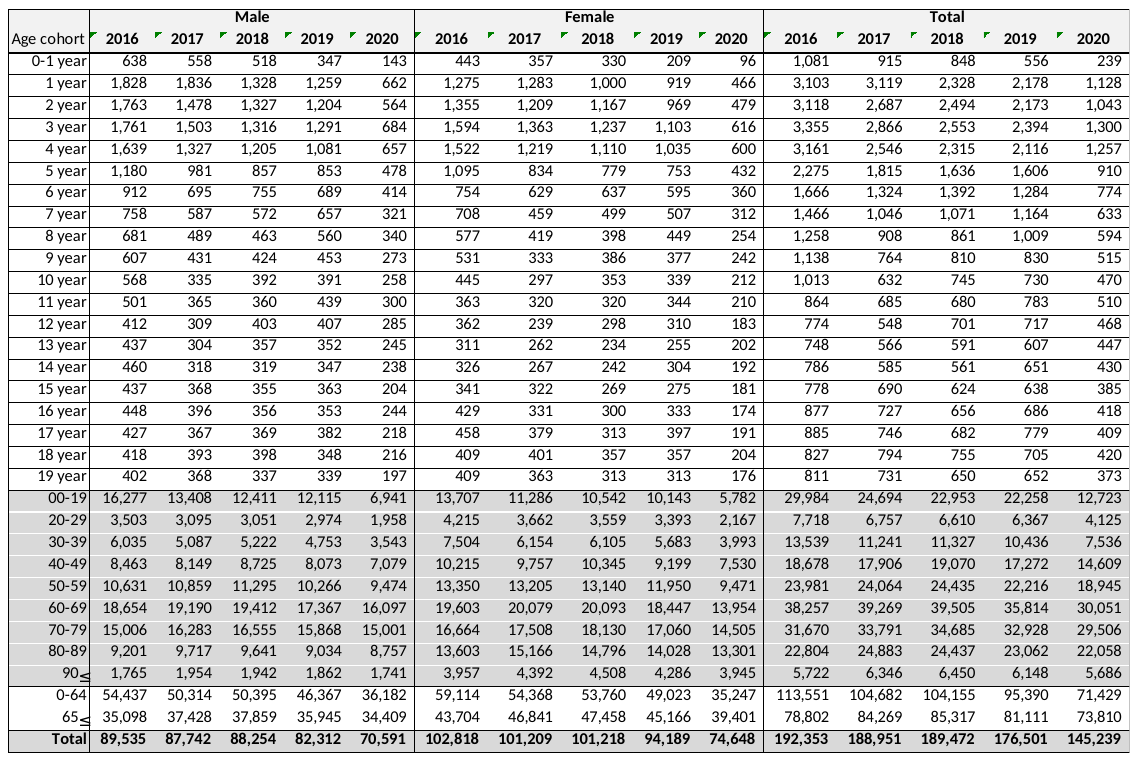

**Supplementary Table 1B** Total annual number of recorded community-acquired pneumonia cases, by study year, age group, and sex (2016-2020)

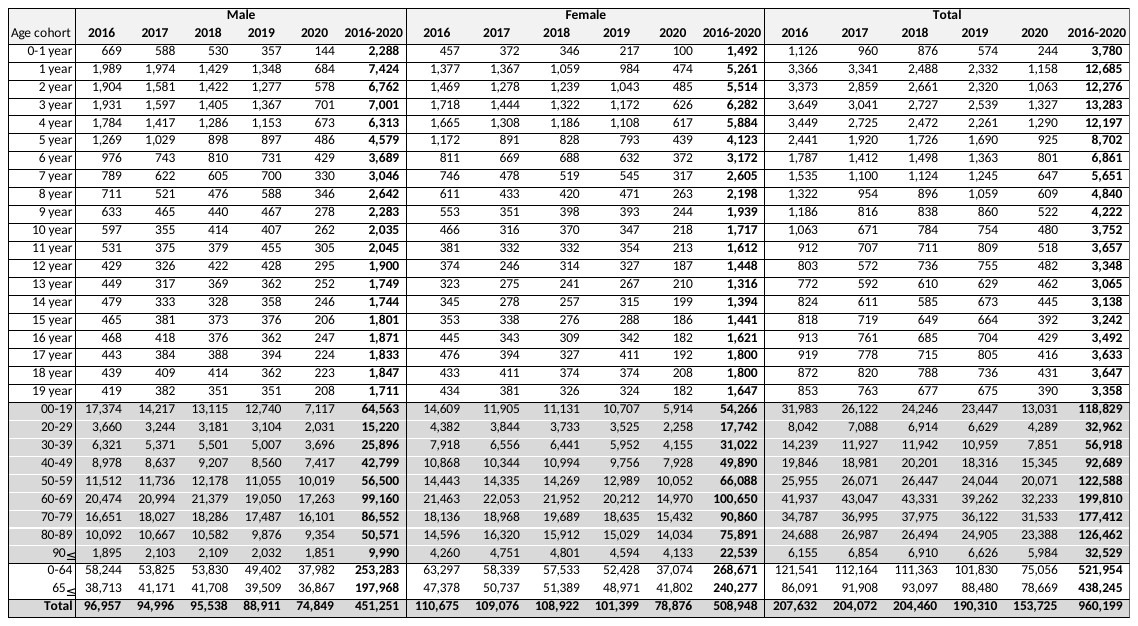

**Supplementary Table 2** Annual population-based rates of diagnosed community-acquired pneumonia per 100,000, by study year, age group, and sex (2016-2020)

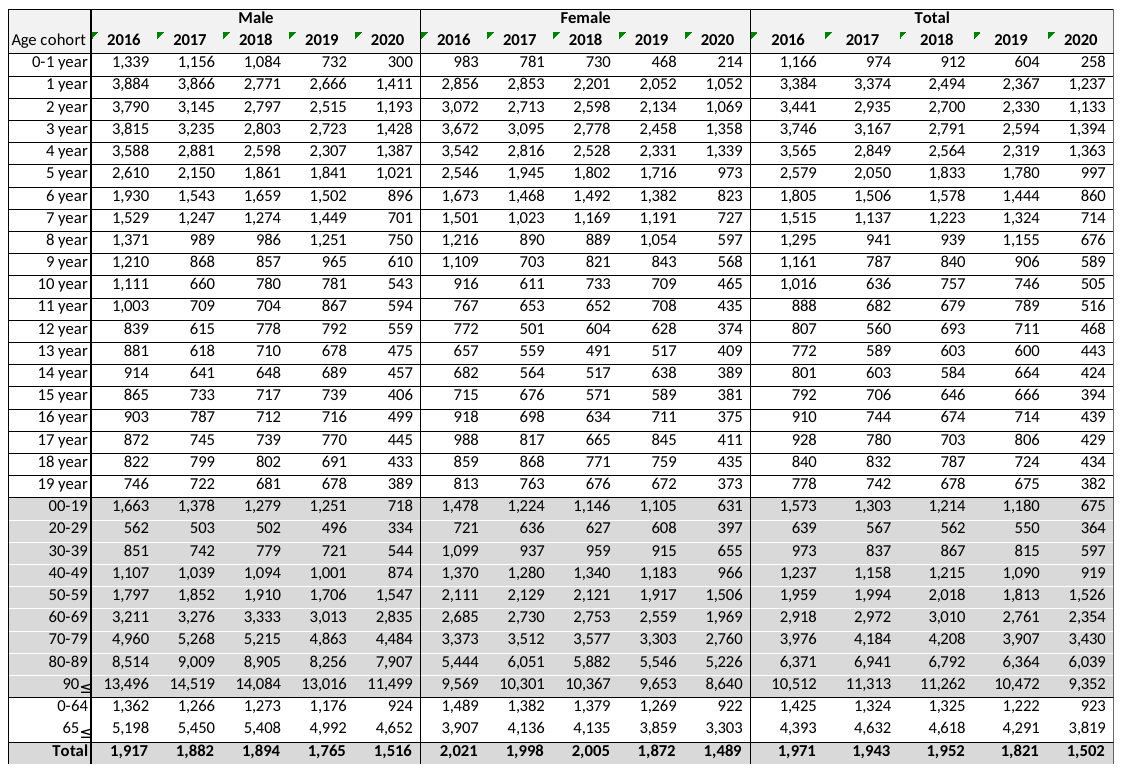
**Supplementary Table 3A.** Number of hospitalizations within 0–15 days after community-acquired pneumonia diagnosis, by study year, age group, and sex (2016–2020).

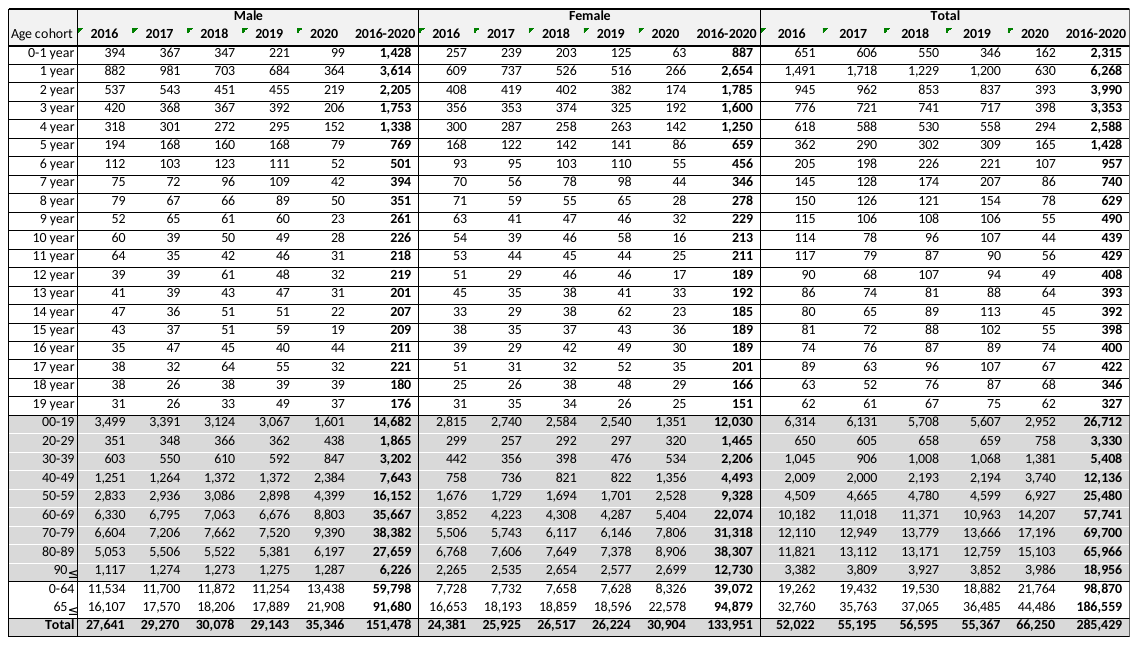

**Supplementary Table 3B.** Proportion of patients hospitalized within 0–15 days after community-acquired pneumonia diagnosis, by study year, age group, and sex (2016–2020).

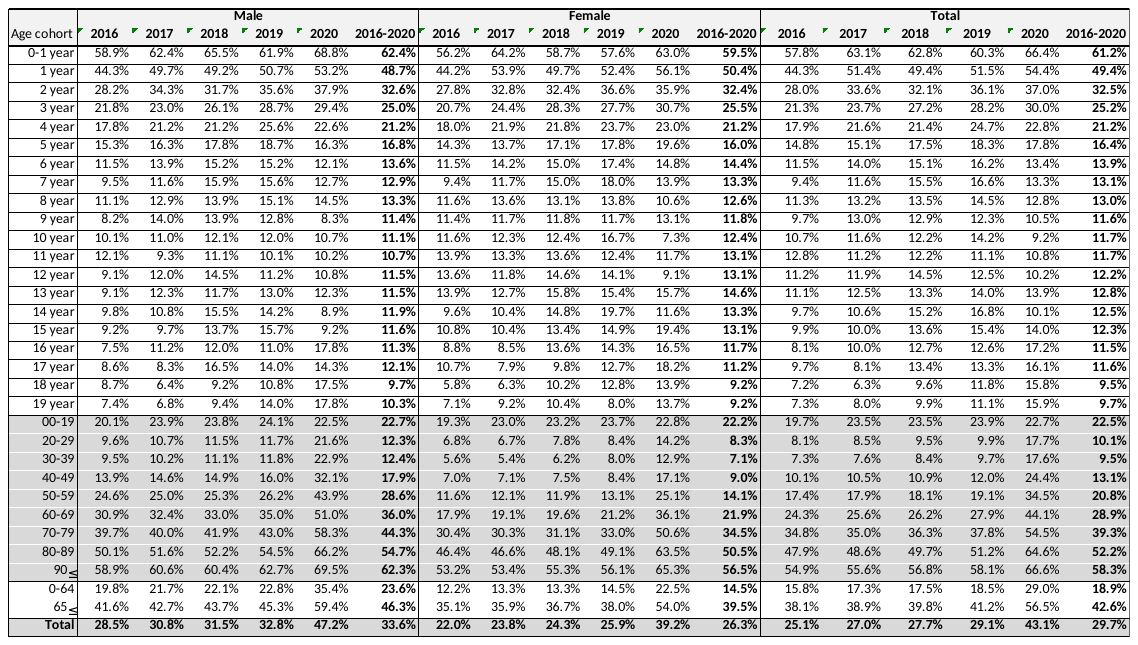

**Supplementary Table 4A** Number of deaths within 30 days of diagnosis among patients with community-acquired pneumonia hospitalized within 0–15 days after diagnosis, by study year, age group, and sex (2016–2020).

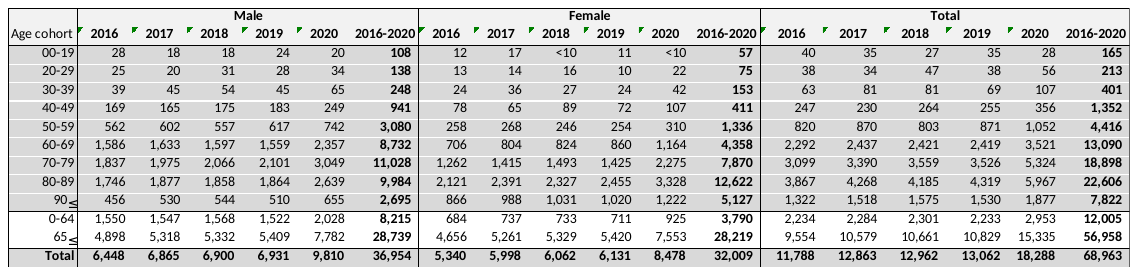

**Supplementary Table 4B** 30-day all-cause mortality among patients with community-acquired pneumonia hospitalized within 0-15 days after diagnosis, by study year, age group, and sex (2016-2020)

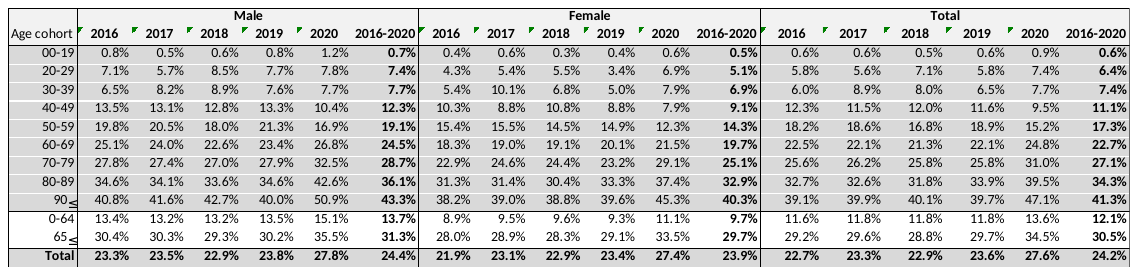
